# Effect of daily human movement on some characteristics of dengue dynamics

**DOI:** 10.1101/2020.09.20.20198093

**Authors:** Mayra R. Tocto-Erazo, Daniel Olmos-Liceaga, Jose A. Montoya-Laos

## Abstract

Human movement is a key factor in infectious diseases spread such as dengue. Here, we explore a mathematical modeling approach based on a system of ordinary differential equations to study the effect of human movement on characteristics of dengue dynamics such as the existence of endemic equilibria, and the start, duration, and amplitude of the outbreak. The model considers that every day is divided into two periods: high-activity and low-activity. Periodic human movement between patches occurs in discrete times. Based on numerical simulations, we show unexpected scenarios such as the disease extinction in regions where the local basic reproductive number is greater than 1. In the same way, we obtain scenarios where outbreaks appear despite the fact that the local basic reproductive numbers in these regions are less than 1 and the outbreak size depends on the length of high-activity and low-activity periods.

## 1. Introduction

Dengue is an endemic disease in many countries around the world, mainly throughout the tropics [1, 2]. It is estimated that there are a total of 3.97 billion people at risk of dengue transmission [3]. Risk levels depend strongly on rainfall, temperature and the degree of urbanization [1]. Human movement is also a key component of the transmission dynamics of many vector-borne diseases [4, 5]. For example, dengue infections has been related to travel to endemic places such as the Caribbean, South America, South-Central Asia, and Southeast Asia [6].

In urban areas, human movement is frequent and extensive but often composed of commuting patterns between homes and places of employment, education or commerce [7]. At this scale, commuting people occurs day-to-day, dominated by daily activities. In a study conducted at two factories in Bandung [8], authors suggest that some people may have acquired the dengue virus at work and not at home. Therefore, local human movement plays an important role in the temporal and spatial spread of the dengue disease.

From the mathematical point of view, the role of the human movement on vector-borne diseases from various perspectives has been studied. In particular, ordinary differential equations have been used to model the human mobility between two or more locations [9]. One approach is the continuous moving of the human population between places [10–15]. Other proposal is the residence time, which represents the proportion of time that human budget their residence across regions [16–19]. However, other approach is to explicitly consider the daily movement of people on the dynamics. This approach has been studied in [20], where the authors formulate a star-network of connections between a central city and peripheral villages. Also, they suppose the commute population is the same every day and the movement period to the central city is half a day. Despite there are studies about the human movement from different approaches, it has been poorly understood.

In this work, our objective is to study the effect of the daily periodic movement on dengue dynamics such as the existence of endemic equilibria, and the start, duration and, amplitude of the outbreak. We formulate a two-patch model based on a system of ordinary differential equations and incorporate human daily movement, where movement takes place at periodic discrete times every day as in [20]. Every day is divided into two periods: low-activity and high-activity, which could represent night and day, respectively. We consider that the low-activity period represents the time interval where humans stay at their residence patch. The movement takes place during the high-activity period in which people commute to school, work or other daily activities; also a high-activity period can be related to extraordinary events where large numbers of humans interact. To study this model, we first analyze the patches separately without considering a piecewise definition in time. Then, based on numerical simulations, we study the complete model to observe some effects of the human periodic movement on the dynamics.

This work is divided in the following sections. The formulation of the model and the analysis of uncoupled patches is given in Section 2. Then, in Section 3, we study the effect of daily human movement on some characteristics of model dynamics based on numerical studies under some scenarios. Finally, conclusions and discussions about our results are presented in Section 4.

## 2. Formulation of model

The classic vector-host mathematical model is given by the following system

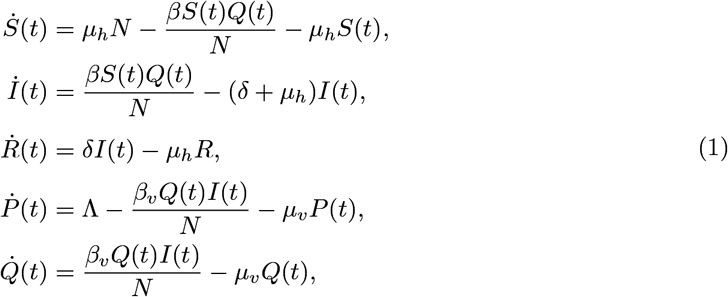

where *S, I* and *R* represent the susceptible, infected and recovered population, respectively, and *P* y *Q* the susceptible and infected mosquito population, respectively.

We include the daily periodic movement between two patches in model (1) as follows. The interval [*t*_*k*_, *t*_*k*+1_) is the time period corresponding to the *k*th day and *T*_*l*_ ∈ (0, 1) the fraction of the day of *low-activity* such that for interval [*t*_*k*_, *t*_*k*_ + *T*_*l*_) we have in each patch only resident population composed of *N*_*i*_ individuals (*i* = 1, 2). Thus, the time interval [*t*_*k*_, *t*_*k*_ + *T*_*l*_) is named the *low-activity* period. For a fixed day *k, α*_*i*_ represents the proportion of the population from patch *i* that moves every day to another patch *j* at time *t*_*k*_ + *T*_*l*_ and returns to patch *i* at time *t*_*k*+1_. Thus, human movement takes place on the time interval [*t*_*k*_ + *T*_*l*_, *t*_*k*+1_) which is named the *high-activity* period.

For the low-activity period, the susceptible, infected and recovered human population from patch *i* are represented by 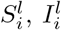 and 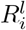 respectively, and the susceptible. The susceptible and infected vector population from patch *i* are represented by *P*_*i*_ and *Q*_*i*_, respectively. On the other hand, for high-activity period, human population from patch *i* is divided into two subpopulations. The first subpopulation is composed of people from patch *i* who do not move to another patch, that is, (1 − *α*_*i*_)*N*_*i*_. This subpopulation is subdivided into susceptible 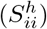, infected 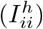 and recovered 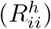. The second subpopulation is composed of residents from patch *j* who move to patch *i, α*_*j*_*N*_*j*_. This subpopulation is subdivided into susceptible 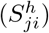, infected 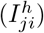 and recovered 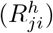. Since we assume that the vector population does not move between patches, susceptible and infected vectors remain represented by *P*_*i*_ and *Q*_*i*_, respectively. Thus, the following equations represent the dynamics of the populations for the low-activity period [*t*_*k*_, *t*_*k*_ + *T*_*l*_):

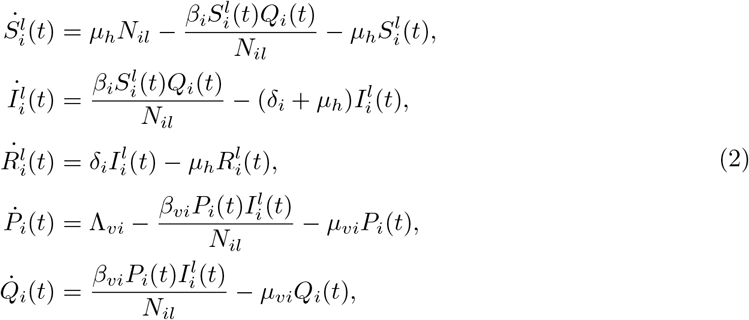

where *N*_*il*_ := *N*_*i*_ and *i* = 1, 2.

For the high-activity period [*t*_*k*_ + *T*_*l*_, *t*_*k*+1_), the set of equations become:

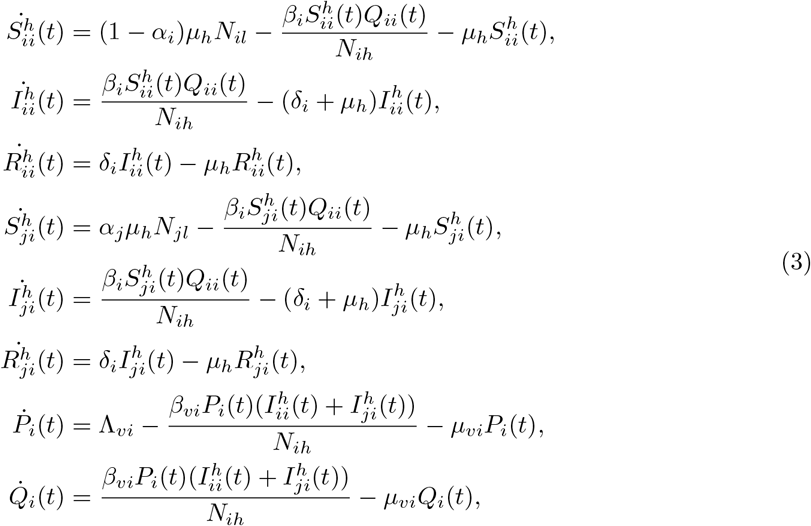

where *N*_*ih*_ := (1 − *α*_*i*_)*N*_*i*_ + *α*_*j*_*N*_*j*_, and *i, j* = 1, 2, *i* ≠ *j*. All model parameters are defined in Table 1.

**Table 1:** Parameter definition of model (2)-(3).

| Parameter | Meaning |
| --- | --- |
| $\alpha_i$ | Proportion of humans from patch $i$ who move to patch $j$ at time $t_k + T_{la}$. |
| $N_i$ | Resident humans of patch $i$. |
| $1/\mu_h$ | Average life time of humans. |
| $1/\mu_{vi}$ | Average life time of mosquitoes in patch $i$. |
| $\beta_i$ | Transmission rate from mosquito to human in patch $i$. |
| $\beta_{vi}$ | Transmission rate from human to mosquito in patch $i$. |
| $1/\delta_i$ | Average recovery time of humans in patch $i$. |
| $\Lambda_{vi}$ | Mosquito recruitment rate in patch $i$. |

We observe that model (2)-(3) can be reduced to uncoupled patches in the form of system (2). This is done by taking *T l* = 1, that is, having only low-activity periods.

In order to study our coupled model (2)-(3), we first make an analysis for each system separately without considering a piecewise definition in time. Then, we focus on understanding the dynamics of the daily human movement.

### 2.1. Uncoupled case

System (2) is positively invariant in 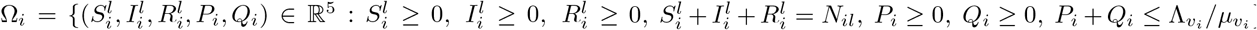 [21]. Then, the disease-free equilibrium of system (2) is given by 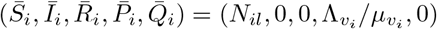 and, using to the next generation matrix approach as in [22], the basic reproductive number (*R*_*il*_) of uncoupled system is given by

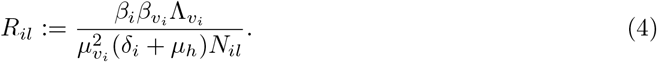

Previous work [21] has shown that if *R*_*il*_ > 1, then there exists an endemic equilibrium 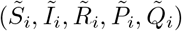, where

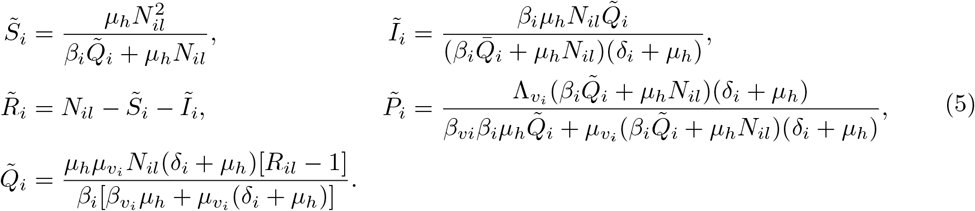

In addition, authors in [21, 23] also have shown that the disease-free equilibrium is globally asymptotically stable (GAS) when *R*_*il*_ < 1, and the endemic equilibrium is GAS when *R*_*il*_ > 1.

For the high-activity period (3), we define 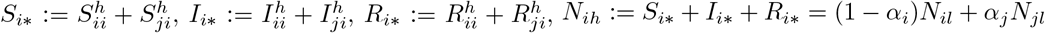. Thus, the dynamics of uncoupled system (3) can be written as:

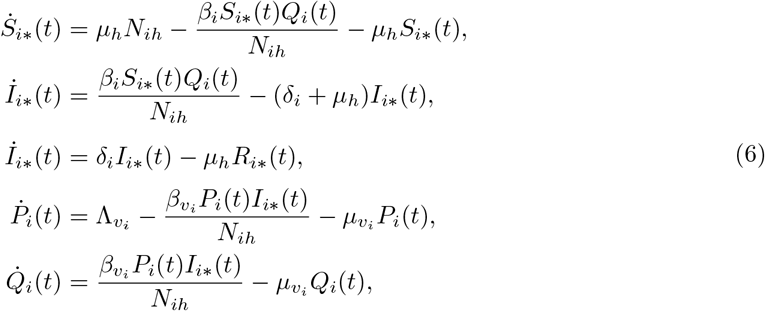

for each *i* = 1, 2.

Since the structure of system (6) is the same as (2), results concerning the stability of the equilibrium points are analogous to system (2). In particular, the disease-free and endemic equilibrium points are given by 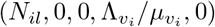 and 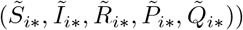, respectively, where

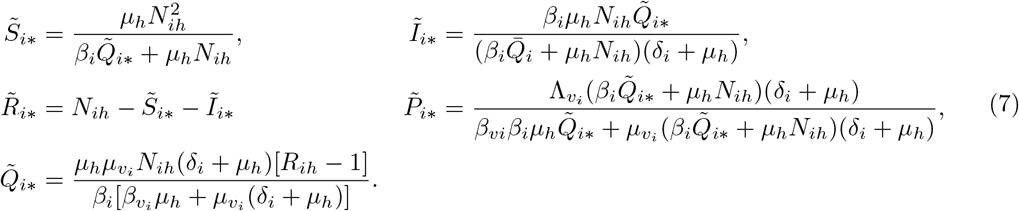

In addition, the basic reproductive number (*R*_*ih*_) for uncoupled system (6) is given by

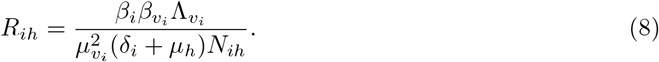

We observe that each *R*_*il*_ and *R*_*ih*_ does not depend on human movement, in this sense, the theoretical results on the existence and stability of the equilibrium points are given for the uncoupled system. However, when we the patches are coupled, these basic reproductive numbers loses meaning and only provide information when the patches are uncoupled. In this case, a global *R*_0_ is not well defined so that we want to give an understanding of what occurs in each patch due to the movement depending on the local basic reproductive numbers.

## 3. Effect of daily human movement on the endemic levels and the outbreaks

In this section, we focus on understanding some effects due to daily human movement on the existence of the endemic equilibrium points and on the outbreaks for coupled model (2)-(3). For this, we proceed to study such effects in three stages. In the first stage, we characterize the local basic reproductive number and the endemic equilibrium values of each patch as a function of the total population size, in order to see how changes in the population affect both, the *R*_0_ and equilibrium point values. In the second stage, we show the changes that the local basic reproductive numbers in each patch may experiment after migration. In the last stage, based on numerical studies, we evidence the effects of daily human movement on some characteristics of the dynamics such as the existence and disappearance of endemic equilibria, duration, size and peak of the outbreak.

### 3.1. Dependence on the basic reproductive number and the endemic equilibria as a function of population size

In general, the basic reproductive number (*R*_0_) and the endemic equilibrium (*I*^*\**^) of model (2)-(3) for a disconnected patch with human population *N* can be written as

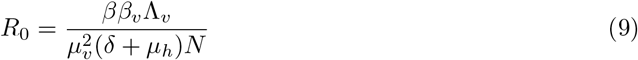

and

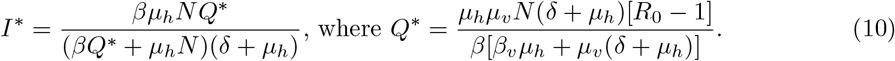

From (9) and (10), we have that *R*_0_ = 1 at a point 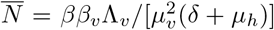, and *I*^*\**^ reaches its maximum at point 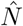 given by

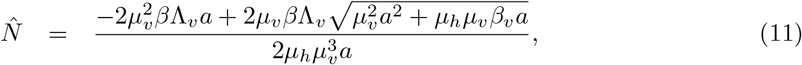

where *a* = *δ*+ *µ*_*h*_. From Figure 1, we observe that a patch with *N* smaller (larger) than 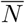 leads to have *R*_0_ > 1 (*R*_0_ < 1). The basic reproductive number is a measure that gives conditions for the existence of endemic equilibria and disease propagation in each patch separately. Thus, *R*_0_ < 1 means that there is no favorable conditions for the disease spread, whereas *R*_0_ > 1 implies that the conditions are favorable for an outbreak to occur in each disconnected patch. In addition, while 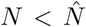, the value of the endemic equilibrium *I*^*\**^ increases as *N* grows up, and decreases when 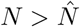.

**Figure 1:**
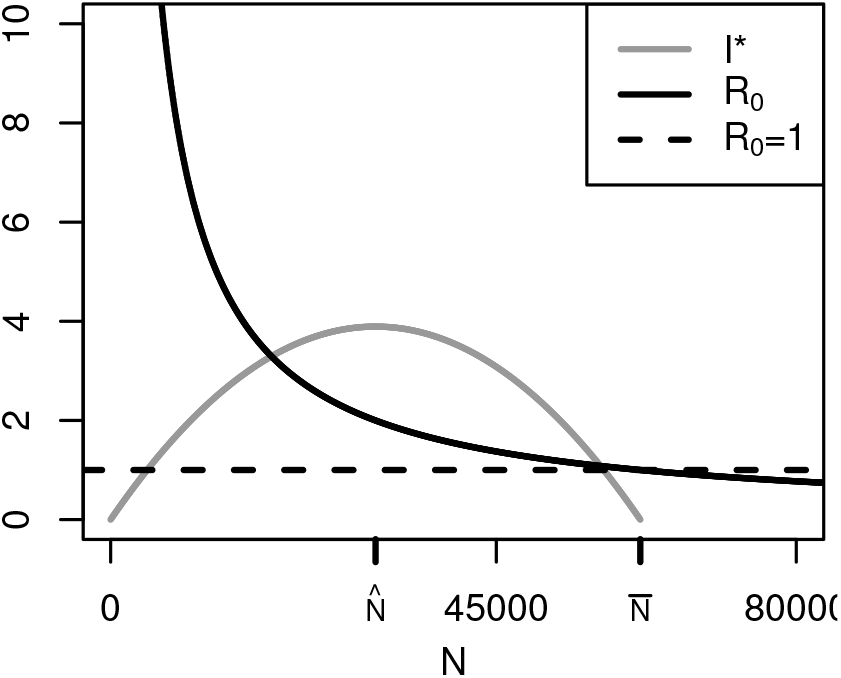
*R*_0_ and *I*^*\**^ versus *N*. Parameter values: Λ_*v*_ = 1200, *β* = 0.25, *β*_*v*_ = 0.15, *µ*_*h*_ = 0.000036, *µ*_*v*_ = 0.0714, and *δ*= 0.1428.

### 3.2. Changes in R_0_ after one migration process

The findings in the previous subsection can be applied to see how the disease propagation conditions change in each patch when there is human migration between them. For this, we define *A* as the net population that move between patches, that is, *A* := |*α*_1_*N*_1*l*_ − *α*_2_*N*_2*l*_|. Table 2 shows a list of possible outcomes of the basic reproductive numbers after the interchange of populations from one patch to another. The results of Table 2 are based on the value of 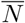 which is the threshold population that generates or not endemic equilibria. The first column of the table shows the value of the basic reproductive number in each patch before migration is considered (*R*_1*l*_ and *R*_2*l*_). The second column shows the possible outcomes after a proportion of humans from patch 1 moves to patch 2, and vice versa (*R*_1*h*_ and *R*_2*h*_). The third column displays the conditions that the populations must satisfy in order for every scenario to occur. The scenarios are used to understand the daily human movement between patches.

**Table 2:** Possible scenarios for *R*_1*h*_ and *R*_2*h*_ after population exchange.

| Scenarios before migration | Scenarios after migration | Conditions |
| --- | --- | --- |
| $R_{1l} < 1$ and $R_{2l} > 1$ | $R_{1h} < 1$ and $R_{2h} < 1$ | $N_{1h} = N_{1l} - A$ and $N_{2h} = N_{2l} + A$<br>where $\bar{N} - N_{2l} < A < N_{1l} - \bar{N}$ |
| | $R_{1h} < 1$ and $R_{2h} > 1$ | $N_{1h} = N_{1l} - A$ and $N_{2h} = N_{2l} + A$<br>where $A < \min \{N_{1l} - \bar{N}, \bar{N} - N_{2l}\}$<br>**or**<br>$N_{1h} = N_{1l} + A$ and $N_{2h} = N_{2l} - A$ |
| | $R_{1h} > 1$ and $R_{2h} < 1$ | $N_{1h} = N_{1l} - A$ and $N_{2h} = N_{2l} + A$<br>where $A > \max \{N_{1l} - \bar{N}, \bar{N} - N_{2l}\}$ |
| | $R_{1h} > 1$ and $R_{2h} > 1$ | $N_{1h} = N_{1l} - A$ and $N_{2h} = N_{2l} + A$<br>where $N_{1l} - \bar{N} < A < \bar{N} - N_{2l}$ |
| $R_{1l} < 1$ and $R_{2l} < 1$ | $R_{1h} < 1$ and $R_{2h} < 1$ | $N_{1h} = N_{1l} - A$ and $N_{2h} = N_{2l} + A$<br>where $A < N_{1l} - \bar{N}$<br>**or**<br>$N_{1h} = N_{1l} + A$ and $N_{2h} = N_{2l} - A$<br>where $A < N_{2l} - \bar{N}$ |
| | $R_{1h} < 1$ and $R_{2h} > 1$ | $N_{1h} = N_{1l} + A$ and $N_{2h} = N_{2l} - A$<br>where $A > N_{2l} - \bar{N}$ |
| | $R_{1h} > 1$ and $R_{2h} < 1$ | $N_{1h} = N_{1l} - A$ and $N_{2h} = N_{2l} + A$<br>where $A > N_{1l} - \bar{N}$ |
| $R_{1l} > 1$ and $R_{2l} > 1$ | $R_{1h} > 1$ and $R_{2h} > 1$ | $N_{1h} = N_{1l} + A$ and $N_{2h} = N_{2l} - A$<br>where $A < \bar{N} - N_{1l}$<br>**or**<br>$N_{1h} = N_{1l} - A$ and $N_{2h} = N_{2l} + A$<br>where $A < \bar{N} - N_{2l}$ |
| | $R_{1h} > 1$ and $R_{2h} < 1$ | $N_{1h} = N_{1l} - A$ and $N_{2h} = N_{2l} + A$<br>where $A > \bar{N} - N_{2l}$ |
| | $R_{1h} < 1$ and $R_{2h} > 1$ | $N_{1h} = N_{1l} + A$ and $N_{2h} = N_{2l} - A$<br>where $A > \bar{N} - N_{1l}$ |

In order to show how the displacement of people from one patch to another may influence the disease propagation conditions, we examine the scenario *R*_1*l*_ < 1 and *R*_2*l*_ > 1, i.e, during the low-activity period, in patch 1, the disease propagation conditions are not favorable, and in patch 2, the conditions are favorable. To this, we consider the following resident populations: *N*_1*l*_ = 90000 and *N*_2*l*_ = 45000 for patch 1 and 2, respectively, and parameter values given in Table 3. Based on the parameter values, we obtain that *R*_1*l*_ = 0.68 and *R*_2*l*_ = 1.37. Figure 2 shows under which conditions *R*_1*h*_ and *R*_2*h*_ are smaller or greater than 1, where the latter results in the existence of endemic equilibria according to *α*_1_ and *α*_2_ values. Note that Figure 2 shows only the first three outcomes for the case *R*_1*l*_ < 1 and *R*_2*l*_ > 1 given by Table 2. Observe that there are no values of *α*_1_ and *α*_2_ where both *R*_1*h*_ and *R*_2*h*_ are simultaneously greater than 1.

**Table 3:** Parameter values for the different scenarios. All parameter values are taken from [24]. Values for Λ_*v*1_ and Λ_*v*2_ are given in this study.

| Parameter | Value |
| --- | --- |
| $\mu_h$ | 0.000036 |
| $\mu_{v1}, \mu_{v2}$ | 0.0714 |
| $\beta_1, \beta_2$ | 0.25 |
| $\beta_{v1}, \beta_{v2}$ | 0.15 |
| $\delta_1, \delta_2$ | 0.1428 |
| $\Lambda_{v1}, \Lambda_{v2}$ | 1200 |

**Figure 2:**
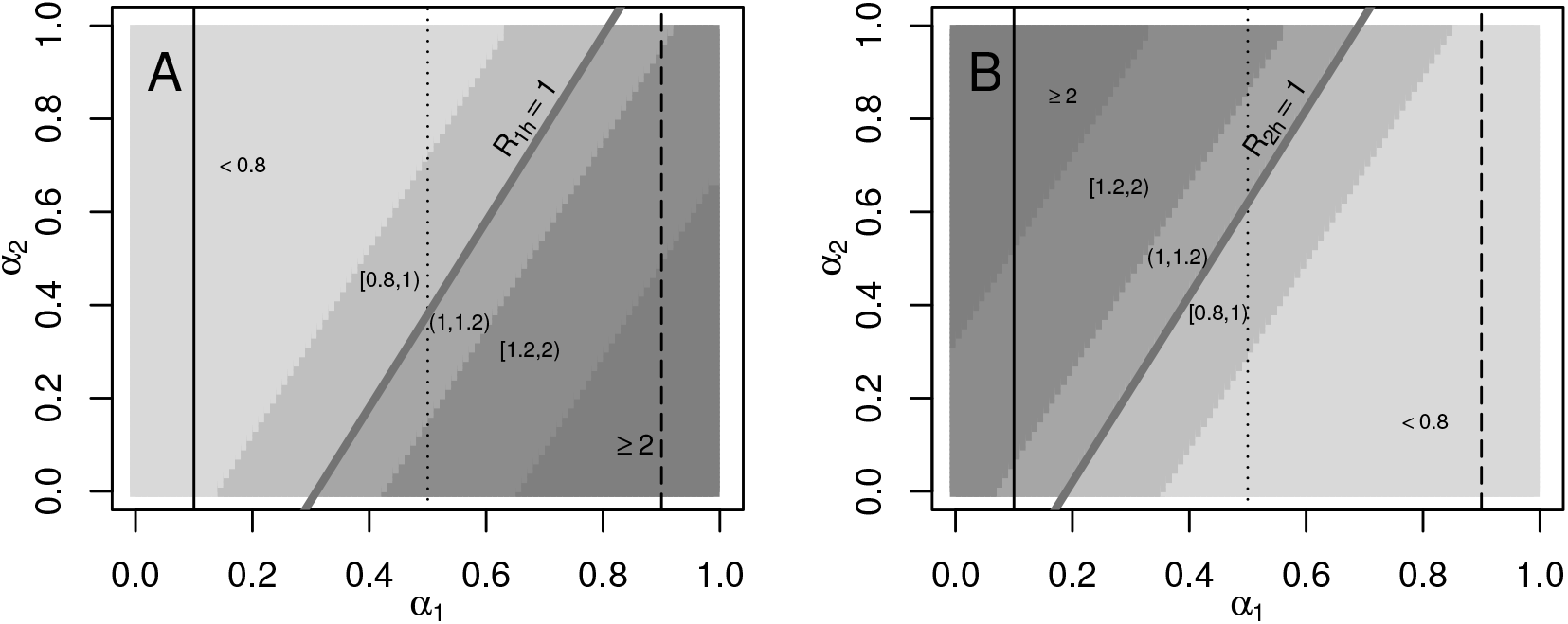
Value regions for *R*_1*h*_ and *R*_2*h*_ according to the *α*_1_ and *α*_2_ values for the scenario *R*_1*l*_ > 1 and *R*_2*l*_ < 1 in (**A**) patch 1 and (**B**) patch 2. The gray thick lines denotes the region where *R*_1*h*_ = 1 (**A**) and *R*_2*h*_ = 1 (**B**). The black lines drawn represent the scenarios that will be studied in the next section.

From now on, to study the effect of daily periodic movement with complete model (2)-(3), we take variables *I*_1_ and *I*_2_ to represent infected residents from patches 1 and 2, respectively. That is,

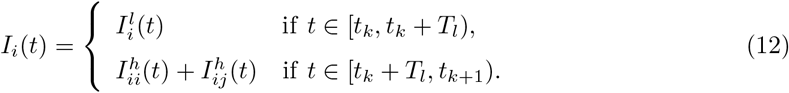

for *i, j* = 1, 2, *i* ≠ *j*. Observe that *I*_*i*_ contabilize the infected individuals from patch *i*, no matter where the disease was acquired.

### 3.3. Numerical studies

In this subsection, we study, by means of numerical simulations, some effects of daily human movement on characteristics of the coupled model solutions, such as the existence of endemic equilibria, and the start, duration, and amplitude of the outbreak.

#### 3.3.1. Disappearance and appearance of endemic equilibria

Here we show the importance of *T*_*l*_, *α*_1_ and *α*_2_ on the existence of endemic equilibria when *R*_1*l*_ < 1 and *R*_2*l*_ > 1. We present numerical simulations for different combinations of these parameters to observe whether or not the existence of endemic equilibria of the uncoupled patches is preserved. Here we study the following cases of the presented scenario in Figure 2: *α*_1_ = 0.1 (black solid line), *α*_1_ = 0.5 (black dotted line), and *α*_1_ = 0.9 (black dashed line), and for every case, we vary *α*_2_ in [0, 1] and *T*_*l*_ in [0.1, 0.9]. Figures 3 to 6 summarize the results of these experiments where we show the values of asymptotic solutions of *I*_1_ and *I*_2_ respect to parameters *α*_2_ and *T*_*l*_. These values will give us an endemic state or a disease-free state.

**Figure 3:**
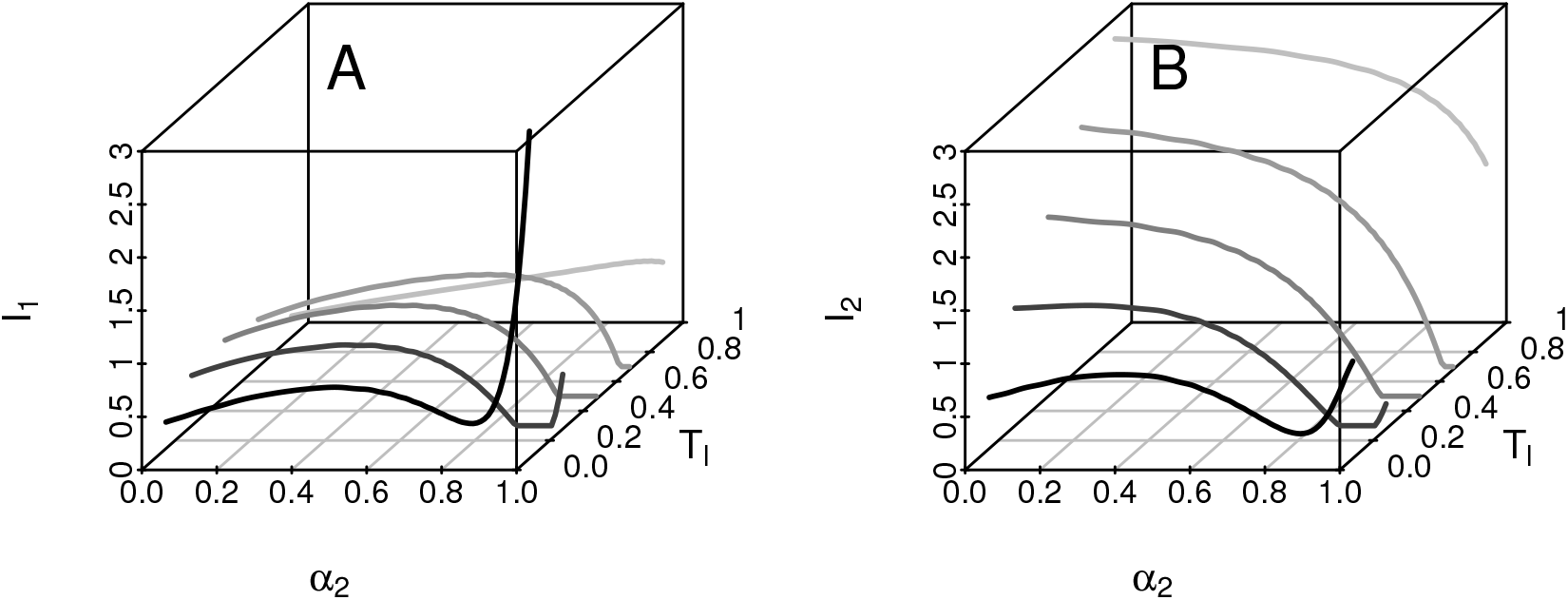
Existence of endemic equilibria for (**A**) *I*_1_ and (**B**) *I*_2_, against *α*_2_ and *T*_*l*_ for case *α*_1_ = 0.1. Population sizes *N*_1*l*_ = 90000 and *N*_2*l*_ = 45000.

##### Case *α*_1_ = 0.1

From Figure 2, theoretically *R*_1*l*_ < 1 and *R*_1*h*_ < 1, that is, there are no favorable conditions at any time during the day in patch 1 for an endemic equilibrium to exist. However, from Figure 3, there exists an endemic equilibrium of patch 1 for almost any combination of *α*_2_ and *T*_*l*_ values. Taking *α*_1_ = 0.1, that is, 10% of individuals from patch 1 move to patch 2, generates endemic levels in patch 1. In general, while the resident people from patch 2 spends more time every day in their own patch, the dynamics are dominated by the theoretical values of *R*_2*l*_ and *R*_2*h*_ which are greater than 1.

From Figure 3, for values of *α*_2_ close to 1 and *T*_*l*_ approximately 0.5, the opposite scenario also occurs. If in a patch there are favorable conditions for the existence of endemic levels all the time (*R*_2*l*_ > 1 and *R*_2*h*_ > 1), we might get no endemic equilibrium in any patch. To get a better understanding of this phenomenon, we examine the behavior of the endemic equilibrium from patches when *α*_2_ = 1.0. Figure 4 shows the existence of endemic equilibria of *I*_1_ and *I*_2_ for *N*_1*l*_ = 90000 with *N*_2*l*_ = 45000 (black solid lines) and *N*_2*l*_ = 18000 (black dashed lines) when *α*_1_ = 0.1 and *α*_2_ = 1.0. For *N*_1*l*_ = 90000 and *N*_2*l*_ = 45000, we have that *R*_1*l*_ = 0.68, *R*_2*l*_ = 1.37, *R*_1*h*_ = 0.49 and *R*_2*h*_ = 6.86. We observe that there is a set of *T*_*l*_ values where the disease disappears in both patches when *N*_2*l*_ = 45000. This phenomenon is explained as follows. Since *α*_1_ = 0.1 and *α*_2_ = 1.0, then, at the beginning of the high-activity period, 10% of the population from patch 1 moves to patch 2 and the whole population from patch 2 moves to patch 1. In the extreme case *T*_*l*_ = 0.9 (the low-activity period is very large), there is an endemic equilibrium in patch 2 due to the fact that almost all the time the population remains in their residence patch and the basic reproductive number (*R*_2*l*_) is greater than 1. In this case, we could approximate the *R*_0_ value of patch 2 by the *R*_0_ value of the disconnected patches. For individuals residing in patch 1, we observe that by taking 10% of individuals from patch 1 who move to patch 2, for a short time period, it is sufficient to generate endemic levels in patch 1 despite theoretically *R*_1*l*_ and *R*_1*h*_ are less than 1. Now, for the extreme case *T*_*l*_ = 0.1 (short low-activity period), the presence of an endemic level in patch 1 is due to the fact that most of the time the 10% of the population that belongs to patch 1, is in patch 2. This 10% carries the endemic levels aquired in patch 2 and take it to patch 1. The endemic levels in patch 2 are due to the presence of endemic levels of mosquitoes that are present in the medium due to the 10% of individuals from patch 1. Finally, for intermediate values of *T*_*l*_, the disease disappears in both patches. For this scenario, both, the resident individuals from patch 2 and the visiting population from patch 1 spend almost the same time in each patch. As the basic reproductive numbers are smaller than 1 in patch 1 and larger than 1 in patch 2, we need to know why the disease cannot be sustained by patch 2. When populations are in patch 2, they do not stay long enough to increase the number of new infected individuals significantly. When individuals move to patch 1, the infective process is much less than in patch 2 (as *R*_1*l*_ = 0.68 < 1 and *R*_1*h*_ = 0.49 < 1) and new infections are imperceptible as the corresponding values of the basic reproductive numbers are very small and not close to 1. The overall effect leads to having a small enough infection rate compared to the disease recovery process and the disease disappears in both patches. However, from Figure 4, this region of disease extinction disappears when *N*_2*l*_ decreases to 18000 (black dashed lines). In this case, we have that *R*_1*h*_ goes up from 0.49 to 0.62 and the region of disease extinction disappear.

**Figure 4:**
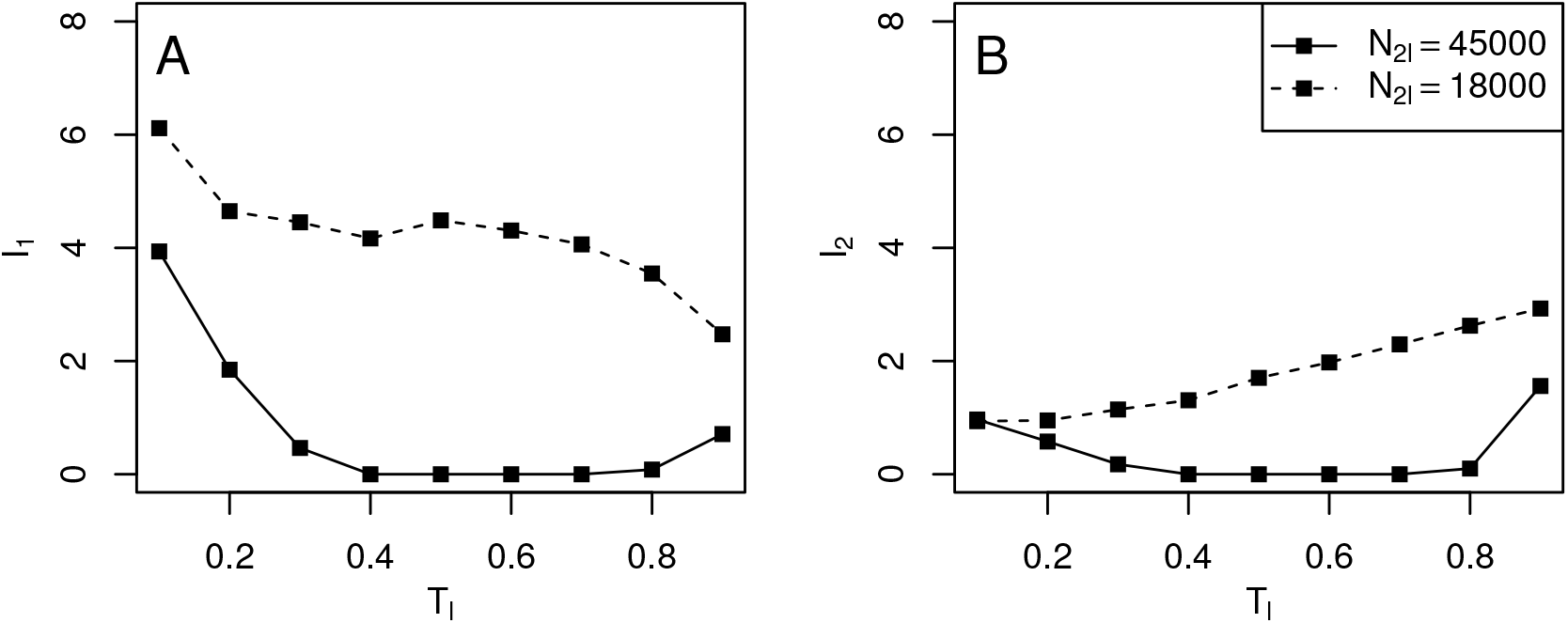
Existence of endemic equilibria for (**A**) *I*_1_ and (**B**) *I*_2_, for *α*_1_ = 0.1 and *α*_2_ = 1.0, and population sizes from patch 2 as *N*_2*l*_ = 45000 (black solid line) and *N*_2*l*_ = 18000 (black dashed line).

##### Cases *α*_1_ = 0.5 and *α*_1_ = 0.9

A similar behavior of existence and non-existence of endemic equilibria arise for these values of *α*_1_ and scenario *R*_1*l*_ < 1 and *R*_2*l*_ > 1.

Figure 5 displays the existence of endemic equilibria of infected residents *I*_1_ and *I*_2_ for *α*_1_ = 0.5. For long periods of low-activity (*T*_*l*_ *≥* 0.7), there are endemic levels in both patches, except for some values of *α*_2_ and *T*_*l*_. In this case (*α*_1_ = 0.5), the region of disease extinction is larger than case *α*_1_ = 0.1. Here, the effect of human movement is more pronounced due to the fact that, for *α*_1_ = 0.5, the values of the basic reproductive numbers for the high-activity period in both patches are in the interval [0.68, 1.37], whereas for *α*_1_ = 0.1, even though *R*_1*h*_ is smaller than 1 (*R*_1*h*_ ∈ [0.49, 0.76]), *R*_2*h*_ takes values in the interval [1.14, 6.54] (see Figure 2).

**Figure 5:**
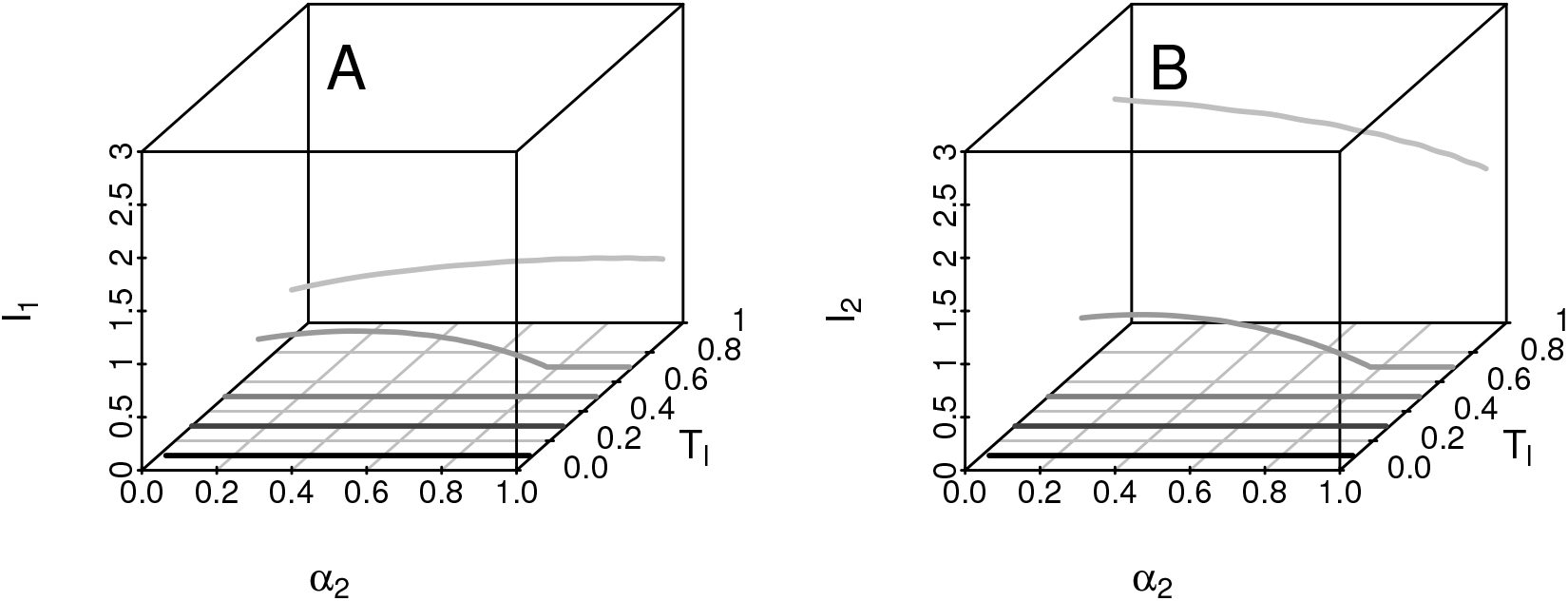
Existence of endemic equilibria for (**A**) *I*_1_ and (**B**) *I*_2_, against *α*_2_ and *T*_*l*_ for case *α*_1_ = 0.5. Population sizes *N*_1*l*_ = 90000 and *N*_2*l*_ = 45000.

Regions of disease extinction can be more complex as is observed in Figure 5, which shows the existence of endemic equilibria of *I*_1_ and *I*_2_ in case *α*_1_ = 0.9. As in the case *α*_1_ = 0.1 and *α*_1_ = 0.5, although there are favorable conditions for the existence of endemic equilibria in one of the patches during the low-activity period, the disease disappears for a set of values of *α*_2_ and *T*_*l*_. For this scenario, the values of *R*_1*h*_ and *R*_2*h*_ are opposite to *R*_1*l*_ and *R*_2*l*_, that is, *R*_1*h*_ > 1 and *R*_2*h*_ < 1. Clearly, depending on the settings of parameters *α*_1_, *α*_2_, *T*_*l*_ and the intensity of the basic reproductive numbers (values of *R*_1*l*_, *R*_2*l*_, *R*_1*h*_ and *R*_2*h*_), different regions of disease extinction can be obtained as observed in Figure 6.

**Figure 6:**
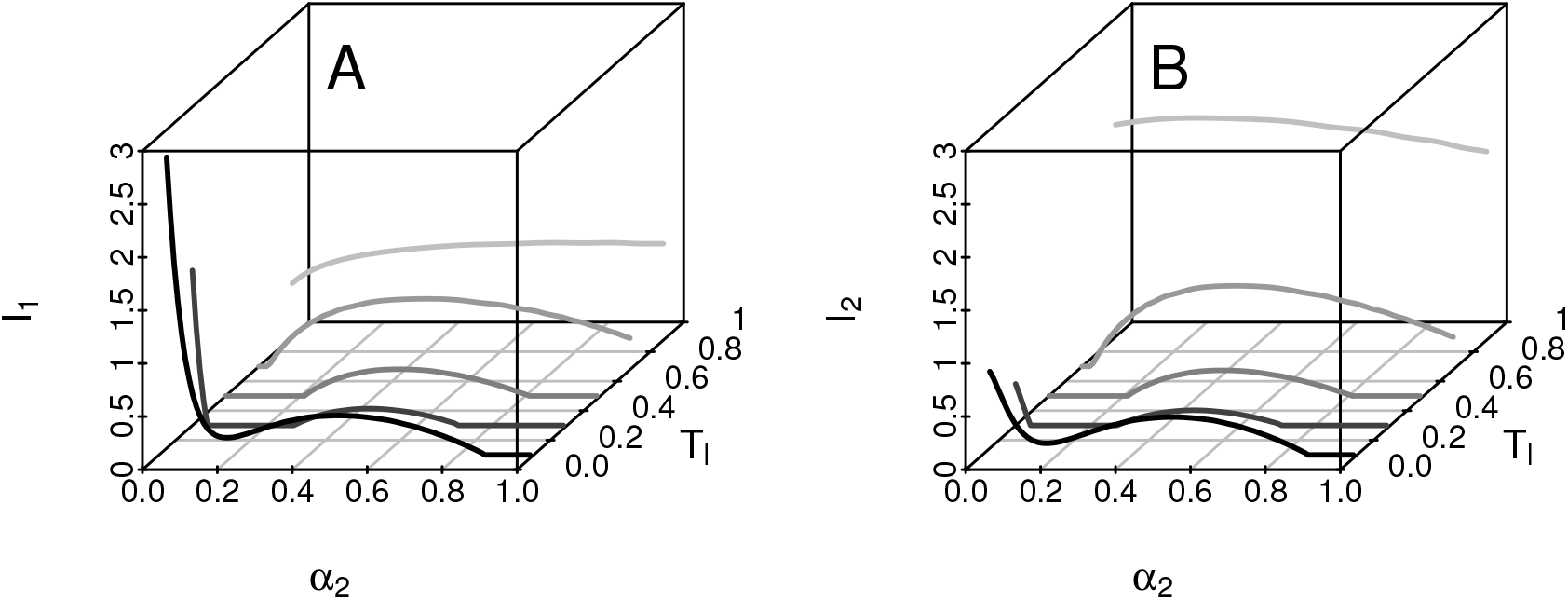
Existence of endemic equilibria for (**A**) *I*_1_ and (**B**) *I*_2_, against *α*_2_ and *T*_*l*_ for case *α*_1_ = 0.9. Population sizes *N*_1*l*_ = 90000 and *N*_2*l*_ = 45000.

#### 3.3.2. Effect on the outbreaks

In this subsection, we present scenarios to observe some effects of the periodic human movement on the outbreak dynamics. The parameter values from Table 3 are used for the numerical simulations.

##### Disappearance of outbreaks

We first explore the scenario given in Subsection 3.3.1, where conditions for the emergence of an outbreak exist only in one of the patches. The purpose is to analyze the complete outbreak in a scenario where the disease disappears.

As we have seen in Figure 3, there are no endemic equilibria in any patch for *α*_2_ values close to 1 and *T*_*l*_ in [0.4, 0.7]. From Figure 7, we observe that there is an oubreak in patch 2 but not in patch 1 when the patches are uncoupled (see dashed lines), which coincides with the fact that *R*_1*l*_ < 1 and *R*_2*l*_ > 1 (*R*_1*l*_ = 0.68 and *R*_2*l*_ = 1.37). If we take *α*_1_ = 0.1 and *α*_2_ = 1.0, implies that *R*_1*h*_ = 0.49 and *R*_2*h*_ = 6.86. Under this scenario, from Figure 7, we notice there are outbreaks in both patches for very long periods of high-activity (*T*_*l*_ = 0.1) and very long periods of low-activity (*T*_*l*_ = 0.9). However, for *T*_*l*_ values in [0.4, 0.7], outbreaks disappear in both patches. That means that this combination of parameters affects not only the existence of endemic equilibria but also the complete existence of the outbreak.

**Figure 7:**
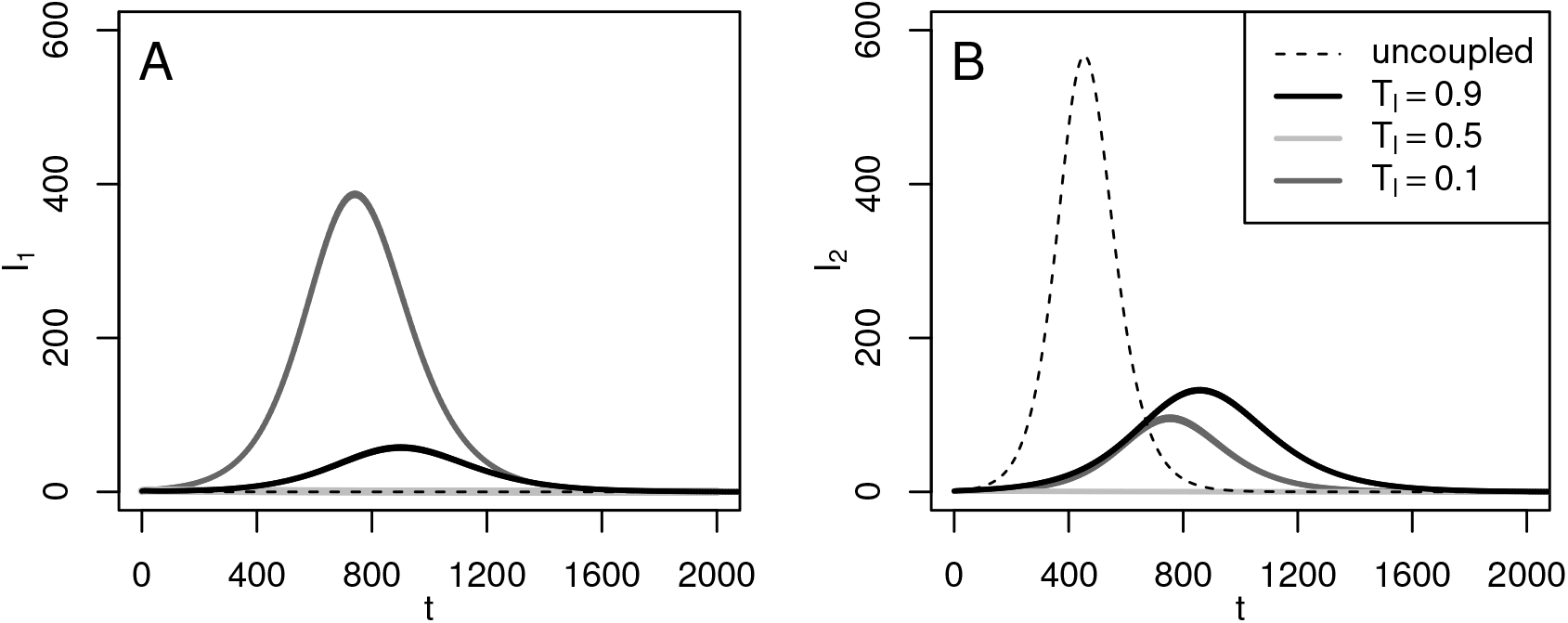
Disappearance of outbreaks. Numerical solutions of infected residents (**A**) *I*_1_ and (**B**) *I*_2_, for *N*_1*l*_ = 90000, *N*_2*l*_ = 45000, *α*_1_ = 0.1 and *α*_2_ = 1.0.

##### Emergence of outbreaks

Here we show the scenario where even though there are no conditions in any of the patches for the existence of an outbreak when patches are uncoupled, there is one due to the human movement. For this, human populations are taken as *N*_1*l*_ = *N*_2*l*_ = 70000, and *α*_1_ = 0 and *α*_2_ = 0.9. For uncoupled patches, there is no outbreak in both patches which coincides with the fact that both *R*_1*l*_ and *R*_2*l*_ are smaller than 1 (*R*_1*l*_ = *R*_2*l*_ = 0.88), and, when there is human movement, theoretically *R*_1*h*_ = 0.46, and *R*_2*h*_ = 8.82. From Figure 8, we observe that an outbreak appears in both patches when *T*_*l*_ = 0.5 approximately and becomes longer as the high-activity period increases. In addition, the time in which the outbreak reaches the highest incidence of cases occurs earlier and is larger as *T*_*l*_ decreases. This phenomenon occurs because as the high-activity period increases, the dynamics of patch 2 are governed by *R*_2*h*_ = 8.82, generating earlier and larger outbreaks in patch 2. For patch 1, in contrast to Figure 7, there is an outbreak in patch 1 despite the fact that there are no favorable conditions for the disease development (*R*_1*l*_ = 0.88 and *R*_1*h*_ = 0.46). However, as outbreaks appear earlier in patch 2 than in patch 1, those infected individuals from patch 2, who move to patch 1 (90% of individuals), interact with mosquitoes from patch 1, generating an outbreak in that patch.

**Figure 8:**
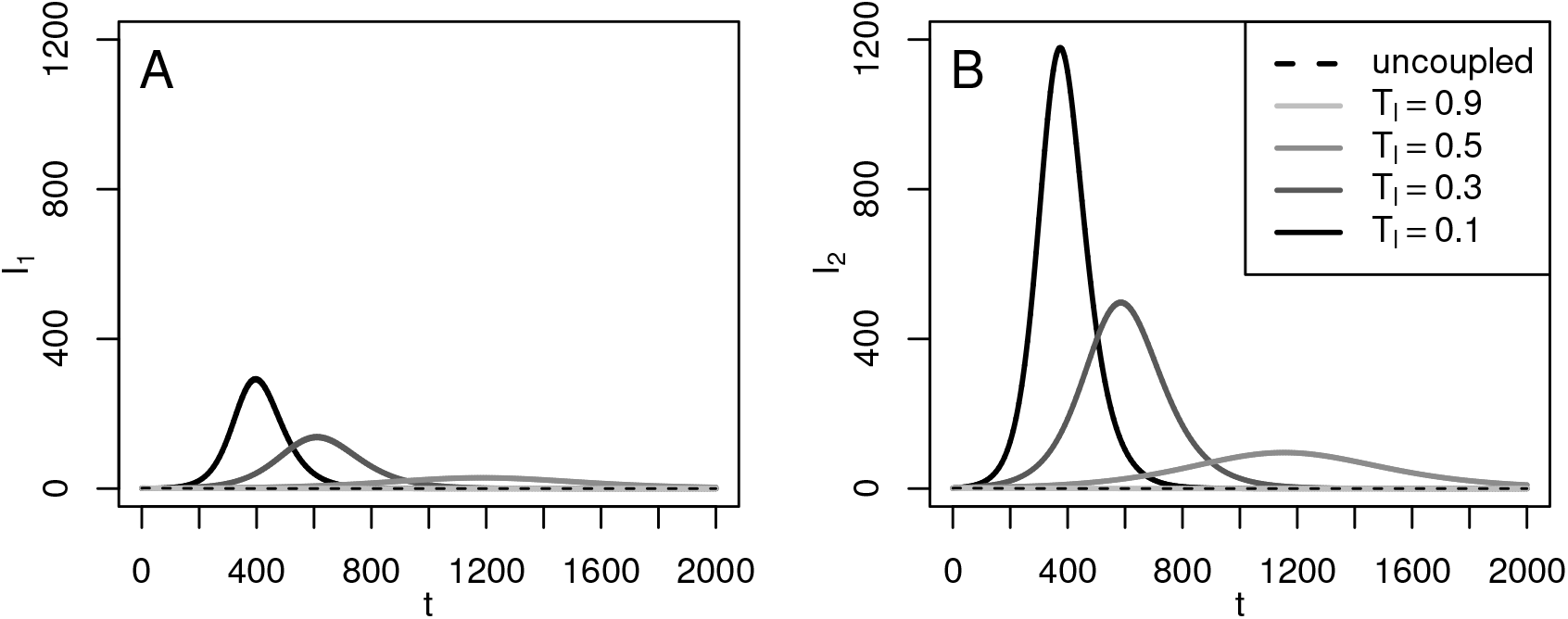
Emergence of outbreaks. Numerical solutions of infected residents (**A**) *I*_1_ and (**B**) *I*_2_, for *N*_1*l*_ = *N*_2*l*_ = 70000, *α*_1_ = 0 and *α*_2_ = 0.9.

##### Delay and advance of outbreaks

To end our study cases, we present scenarios where delay and advance of outbreaks are observed when patches separately have conditions for the existence of outbreaks.

We first take *N*_1*l*_ = 50000, *N*_2*l*_ = 15000, *α*_1_ = 0.5 and *α*_2_ = 0.1. For these values, we obtain *R*_1*l*_ = 1.23, *R*_2*l*_ = 4.11, *R*_1*h*_ = 2.33 and *R*_2*h*_ = 1.60. In Figure 9, we notice that if the patches are uncoupled (black dashed lines), the dynamics of both patches are governed by the *R*_1*l*_ and *R*_2*l*_ values. In this case, the maximum incidence of cases in patch 2 is greater than in patch 1, which coincides with the fact that *R*_2*l*_ is much larger than *R*_1*l*_. Compared to the dynamics of the decoupled patches, the outbreaks for *T*_*l*_ = 0.98 occur earlier in patch 1, and the one in patch 2 remains practically the same. In this case, the temporal dynamics of the uncoupled system are inherited, that is, although the behavior of the outbreak in patch 1 is preserved, this outbreak is advanced because patch 2 has a high incidence of cases. As the high-activity period increases, the maximum incidence in patch 1 goes up and the one in patch 2 decreases. In addition, the outbreak in patch 2 is delayed as *T*_*l*_ goes from 0.98 to 0.02. Clearly, these effects are due to *R*_1*l*_ < *R*_2*l*_ for the low-activity period, but during the high-activity period, the intensity of the basic reproductive numbers is inverted, that is, *R*_1*h*_ > *R*_2*h*_.

**Figure 9:**
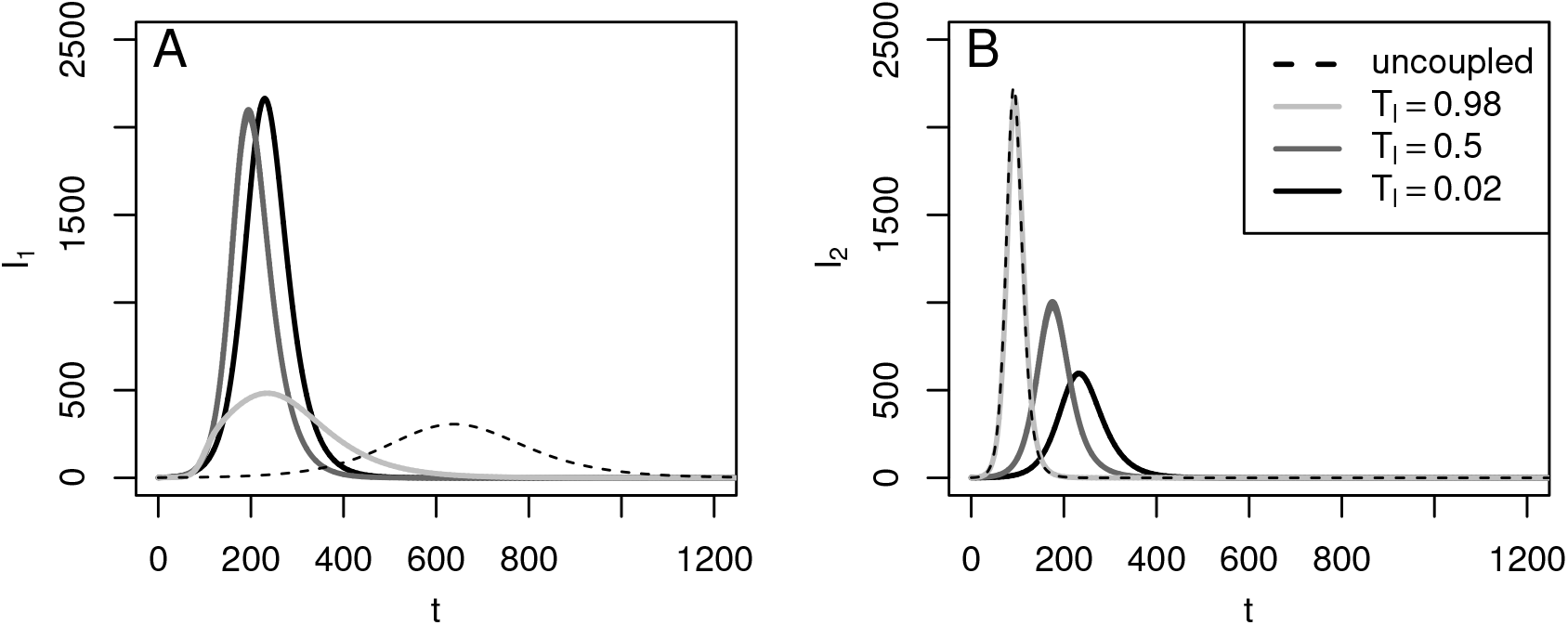
Delay and advance of the outbreaks. Numerical solutions of infected residents (**A**) *I*_1_ and (**B**) *I*_2_, for *N*_1*l*_ = 50000, *N*_2*l*_ = 15000, *α*_1_ = 0.5 and *α*_2_ = 0.1.

Finally, we set *N*_1*l*_ = *N*_2*l*_ = 45000, *α*_1_ = 0.2 and *α*_2_ = 0.8. Thus, we have that *R*_1*l*_ = *R*_2*l*_ = 1.37, *R*_1*h*_ = 0.85 and *R*_2*h*_ = 3.43. In Figure 10, we observe that there are outbreaks when patches are uncoupled (black dashed lines) and these appear earlier when the coupled model is taken into account. Also, the maximum incidence of cases increase as the high-activity period gets longer. In general, this behavior is observed for different settings of *α*_1_ and *α*_2_. To get a better understanding of outbreaks behavior, we analyse how the *R*_1*h*_ and *R*_2*h*_ values change according to proportions of people who move between patches (*α*_1_ and *α*_2_). In fact, since the conditions of disease spread are identical in both patches, this scenario can be studied directly considering only the net population that moved between patches (*A*), defined in Subsection 3.2. For this, we assume, without loss of generality, *R*_1*h*_ < *R*_2*h*_. From Figure 11, we have that while *R*_1*h*_ decreases, *R*_2*h*_ take very large values. In fact, *R*_1*h*_ ∈ [0.68, 1.37], while *R*_2*h*_ can be greater than 20. That is, while *R*_2*h*_ take values very high and *R*_1*h*_ is at least 0.68, the outbreaks appear earlier and the maximum incidence of cases increase.

**Figure 10:**
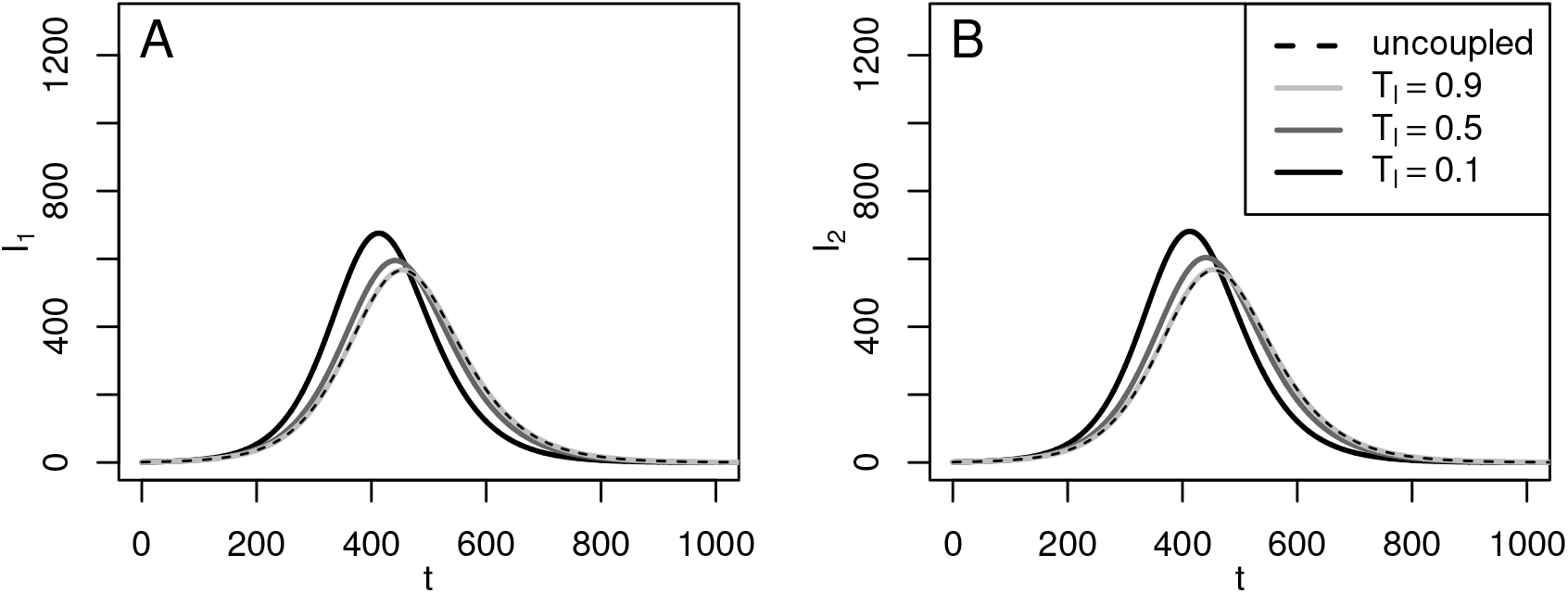
Advance of the outbreaks. Numerical solutions for infected residents (**A**) *I*_1_ and (**B**) *I*_2_, for *N*_1_ = *N*_2_ = 45000, *α*_1_ = 0.2 and *α*_2_ = 0.8.

**Figure 11:**
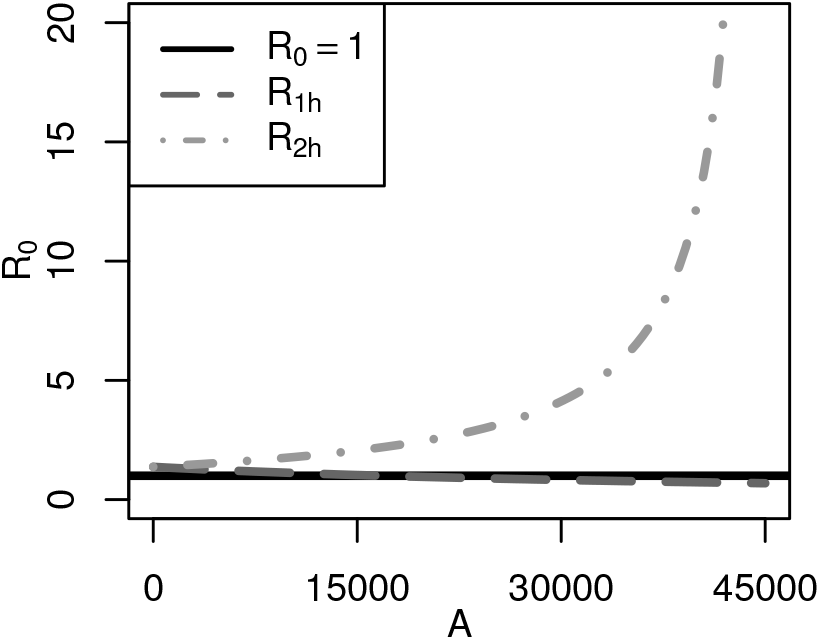
Values of *R*_1*h*_ and *R*_2*h*_ according to the net population that moves between patches (*A*)

## 4. Conclusion and discussion

In this work, our goal was to investigate how the daily human movement affects some characteristics of dengue dynamics based on a two-patch model. The model assumed that the patches are connected by the periodic human movement at discrete times. Given the complexity of the model dynamics, an explicit expression could not be found for the basic reproductive number and the endemic equilibrium points. However, knowing the structure and stability of the equilibrium points and the basic reproductive numbers of the uncoupled system have been useful to determine when there could be favorable or unfavorable conditions for the existence of endemic equilibrium points and outbreaks for the complete model.

This work studies the effects of daily commuters on the disease dynamics under a little-explored approach, different from what is traditionally applied to multi-patch models. We believe that modeling the disease spread where the division of more than one region is clearly defined, needs to be analyzed with a more complete view. Thus, this approach is useful when there are well-defined regions where there is daily human movement between them. Moreover, mixing information from different regions to model it as a single region through a one-patch model without considering movement might give rise to different dynamics than those found in this work. In addition, under this approach, it is possible to recognize where individuals became infected. This fact is important because before applying control measures against possible outbreaks, we should recognize if the cases were imported or autochthonous.

Our scenarios focused on understanding only the effect of human movement on endemic disease levels and on the outbreak dynamics. Some cases of interest were, for example,that although there are regions with disease propagation conditions, the disease do not necessarily subsist. In addition, there could be regions without favorable conditions for the development of the disease, but human movement might lead to the appearance of outbreaks as seen in [25], where the main carriers of the disease between patches is cattle. However, the advantage of the scenarios presented in this work is that it is possible to have a better biological description of the phenomenon.

From Figures 3 to 6, the region of disease extinction varies greatly. We have observed that these regions become larger when the basic reproductive numbers of the uncoupled patches are relatively close to 1. Thus, this fact is dependent on the size of interacting populations and the time spent by the populations in their residence patch. These results are not intuitive and might have not been observed unless the movement between two patches is considered.

Other different scenarios might occur if we assume that the patches have different propagation intensity not related to humans. For example, regions with different sanitary measures or with abundant vegetation could lead to different rates of transmission from humans to mosquitoes or vice versa, mosquito mortality rate, and mosquito recruitment rate. In fact, Table 2 could be generalized considering that the parameters values are not the same in both patches. Also, the model can be extended to a network of patches where individuals from each patch spend different high-activity and low-activity periods in neighboring patches. This extended model might give rise other effects not reported in this work.

Our approach can be useful not only for vector-borne diseases such as zika or chikungunya but also for those with direct transmission such as SARS and COVID-19, diseases which might generate pandemics due to human movement. In this respect, an infected human might be exposed to different populations during its complete period of infection, leading to a more complex understanding of the basic reproductive number and the disease dynamics. Moreover, to obtain a generalization of the basic reproductive number for our complete model might be useful to establish control policies that consider the human movement.

Finally, although the study was mostly computational, it was quite complex. This is due to the number of parameters involved in the model dynamics such as the population sizes of both patches, the proportion of people moving between patches and the time period that individuals spend in their residence patch. Therefore, it is not easy to have a complete theoretical understanding of a system of this nature; however, it was useful to know some properties of the uncoupled model dynamics.

## Data Availability

None.

## References

[1] S. Bhatt, P. W. Gething, O. J. Brady, J. P. Messina, A. W. Farlow, C. L. Moyes, J. M. Drake, J. S. Brownstein, A. G. Hoen, O. Sankoh, M. F. Myers, D. B. George, T. Jaenisch, G. R. W. Wint, C. P. Simmons, T. W. Scott, J. J. Farrar, S. I. Hay, The global distribution and burden of dengue, Nature 496 (2013) 504–507. doi:10.1038/nature12060.

[2] WHO, Dengue and severe dengue, 2019. URL: https://www.who.int/news-room/fact-sheets/detail/dengue-and-severe-dengue.

[3] O. J. Brady, P. W. Gething, S. Bhatt, J. P. Messina, J. S. Brownstein, A. G. Hoen, C. L. Moyes, A. W. Farlow, T. W. Scott, S. I. Hay, Refining the Global Spatial Limits of Dengue Virus Transmission by Evidence-Based Consensus, PLoS Neglected Tropical Diseases 6 (2012). doi:10.1371/journal.pntd.0001760.

[4] S. T. Stoddard, A. C. Morrison, G. M. Vazquez-prokopec, V. P. Soldan, J. Tadeusz, U. Kitron, J. P. Elder, T. W. Scott, The Role of Human Movement in the Transmission of Vector-Borne Pathogens, PLoS neglected tropical diseases 3 (2009). doi:10.1371/journal.pntd.0000481.

[5] S. T. Stoddard, B. M. Forshey, A. C. Morrison, V. A. Paz-soldan, G. M. Vazquez, House-to-house human movement drives dengue virus transmission, Proceedings of the National Academy of Sciences of the United States of America 110 (2013) 994–999. doi:10.1073/pnas.1213349110.

[6] H. P. Mohammed, M. M. Ramos, A. Rivera, M. Johansson, J. L. Muñoz-Jordan, W. Sun, K. M. Tomashek, Travel-associated dengue infections in the United States, 1996 to 2005, Journal of Travel Medicine 17 (2010) 8–14. doi:10.1111/j.1708-8305.2009.00374.x.

[7] B. Adams, D. D. Kapan, Man bites mosquito: Understanding the contribution of human movement to vector-borne disease dynamics, PLoS ONE 4 (2009). doi:10.1371/journal.pone.0006763.

[8] K. R. Porter, C. G. Beckett, H. Kosasih, R. I. Tan, B. Alisjahbana, P. Irani, F. Rudiman, S. Widjaja, E. Listiyaningsih, Epidemiology of dengue and dengue hemorrhagic fever in a cohort of adults living in Bandung, West Java, Indonesia, The American Journal of Tropical Medicine and Hygiene 72 (2005) 60–66. doi:10.4269/ajtmh.2005.72.60.

[9] C. Cosner, Models for the effects of host movement in vector-borne disease systems, Mathematical Biosciences 270 (2015) 192–197. doi:10.1016/j.mbs.2015.06.015.

[10] M. A. Aziz-Alaoui, S. Gakkhar, B. Ambrosio, A. Mishra, A network model for control of dengue epidemic using sterile insect technique, Mathematical Biosciences and Engineering 15 (2017) 441–460. doi:10.3934/mbe.2018020.

[11] A. Mishra, S. Gakkhar, Non-linear Dynamics of Two-Patch Model Incorporating Secondary Dengue Infection, International Journal of Applied and Computational Mathematics 4 (2018). doi:10.1007/s40819-017-0460-z.

[12] R. W. S. Hendron, M. B. Bonsall, The interplay of vaccination and vector control on small dengue networks, Journal of Theoretical Biology 407 (2016) 349–361. doi:10.1016/j.jtbi.2016.07.034.

[13] Y. Xiao, X. Zou, Transmission dynamics for vector-borne diseases in a patchy environment, Journal of Mathematical Biology 69 (2014) 113–146. doi:10.1007/s00285-013-0695-1.

[14] J. Arino, P. Van Den Driessche, A multi-city epidemic model, Mathematical Population Studies 10 (2003) 175–193. doi:10.1080/08898480306720.

[15] G. R. Phaijoo, D. B. Gurung, Mathematical Study of Dengue Disease Transmission in Multi-Patch Environment, Applied Mathematics (2016) 1521–1533. doi:10.4236/am.2016.714132.

[16] S. Lee, C. Castillo-Chavez, The role of residence times in two-patch dengue transmission dynamics and optimal strategies, Journal of Theoretical Biology 374 (2015) 152–164. doi:10.1016/j.jtbi.2015.03.005.

[17] D. Bichara, C. Castillo-Chavez, Vector-borne diseases models with residence times - A Lagrangian perspective, Mathematical Biosciences 281 (2016) 128–138. doi:10.1016/j.mbs.2016.09.006.

[18] J. E. Kim, H. Lee, C. H. Lee, S. Lee, Assessment of optimal strategies in a two-patch dengue transmission model with seasonality, PLoS ONE 12 (2017) 1–21. doi:10.1371/journal.pone.0173673.

[19] E. Barrios, S. Lee, O. Vasilieva, Assessing the effects of daily commuting in two-patch dengue dynamics: A case study of Cali, Colombia, Journal of Theoretical Biology 453 (2018) 14–39. doi:10.1016/j.jtbi.2018.05.015.

[20] E. A. Mpolya, K. Yashima, H. Ohtsuki, A. Sasaki, Epidemic dynamics of a vector-borne disease on a villages-and-city star network with commuters, Journal of Theoretical Biology 343 (2014) 120–126. doi:10.1016/j.jtbi.2013.11.024.

[21] L. Esteva, C. Vargas, Analysis of a dengue disease transmission model, Matematical Biosciences 150 (1998) 131–151. doi:10.1016/S0025-5564(98)10003-2.

[22] F. Brauer, C. Castillo-chavez, A. Mubayi, S. Towers, Some models for epidemics of vector-transmitted diseases, Infectious Disease Modelling (2016) 1–9. URL: http://dx.doi.org/10.1016/j.idm.2016.08.001. doi:10.1016/j.idm.2016.08.001.

[23] M. O. Souza, Multiscale analysis for a vector-borne epidemic model, Journal of Mathematical Biology 68 (2014) 1269–1293. doi:10.1007/s00285-013-0666-6. arXiv:1108.1999.

[24] M. Andraud, N. Hens, C. Marais, P. Beutels, Dynamic epidemiological models for dengue transmission: a systematic review of structural approaches., PloS one 7 (2012) e49085. doi:10.1371/journal.pone.0049085.

[25] M. A. Acuña Zegarra, D. Olmos-Liceaga, J. X. Velasco-Hernández, The role of animal grazing in the spread of chagas disease, Journal of Theoretical Biology 457 (2018) 19–28. doi:10.1016/j.jtbi.2018.08.025.

